# Limited induction of lung-resident memory T cell responses against SARS-CoV-2 by mRNA vaccination

**DOI:** 10.1101/2022.05.25.22275300

**Authors:** Daan K.J. Pieren, Sebastián G. Kuguel, Joel Rosado, Alba G. Robles, Joan Rey-Cano, Cristina Mancebo, Juliana Esperalba, Vicenç Falcó, María J. Buzón, Meritxell Genescà

## Abstract

Resident memory T cells (T_RM_) present at the respiratory tract may be essential to enhance early SARS-CoV-2 viral clearance, thus limiting viral infection and disease. While long-term antigen (Ag)-specific T_RM_ are detectable beyond 11 months in the lung of convalescent COVID-19 patients after mild and severe infection, it is unknown if mRNA vaccination encoding for the SARS-CoV-2 S-protein can induce this frontline protection. We found that the frequency of CD4^+^ T cells secreting interferon (IFN)γ in response to S-peptides was variable but overall similar in the lung of mRNA-vaccinated patients compared to convalescent-infected patients. However, in vaccinated patients, lung responses presented less frequently a T_RM_ phenotype compared to convalescent infected individuals and polyfunctional CD107a^+^ IFNγ^+^ T_RM_ were virtually absent. Thus, a robust and broad T_RM_ response established in convalescent-infected individuals may be advantageous in limiting disease if the virus is not blocked by initial mechanisms of protection, such as neutralization. Still, mRNA vaccines might induce responses within the lung parenchyma, potentially contributing to the overall disease control.

## Introduction

The COVID-19 pandemic continues, and many countries face multiple resurgences. While vaccines to limit SARS-CoV-2 infection rapidly emerged providing high protection from COVID-19, more insight into the mechanisms of protection induced by available vaccines is still needed. The level of vaccine-induced neutralizing antibodies has been shown to correlate with protection from symptomatic infection; however, predicted antibody-mediated vaccine efficacy declines over time ^1^. Moreover, many viral variants of concern (VOC) can significantly evade humoral immunity, yet cellular responses induced by vaccines show strong cross-protection against these variants ^2, 3^, supporting the idea that cellular responses largely contribute to disease control ^4^. In fact, preexisting cross-reactive memory T cells and early Nucleocapsid (N) responses against coronaviruses are associated with protection from SARS-CoV-2 infection ^5, 6^.

Further, SARS-CoV-2 infection induces robust cellular immunity detectable beyond 10 months after infection in peripheral blood ^7^, and as T_RM_ in the lung ^8^, and the number of SARS-CoV-2-specific T_RM_ in the lung correlates with clinical protection ^9^. Vaccination against SARS-CoV-2 using BTN162b2 (Pfizer/BioNTech) and mRNA-1273 (Moderna) vaccines has been reported to induce CD4^+^ and CD8^+^ T-cell responses in peripheral blood ^10, 11^. Moreover, the IFNγ T-cell response to SARS-CoV-2 S-peptides, one of the main antiviral factors measured as a readout, further increased after boosting ^11^. However, current studies only address vaccine-induced SARS-CoV-2 specific T-cell responses in peripheral blood and whether mRNA vaccines also elicit SARS-CoV-2-specific long-term T_RM_ cells in the lung remains to be established.

To this end, we determined the presence of SARS-CoV-2-specific CD4^+^ and CD8^+^ T cells in 26 paired peripheral blood and lung cross-sectional samples from: I.) uninfected unvaccinated individuals (Ctrl, n=5), II.) unvaccinated long-term SARS-CoV-2 convalescent individuals (Inf, n=9, convalescent for a median of 304 days [183-320 IQR]), III.) uninfected and long-term two-dose vaccinated individuals (Vx2, n=7, a median of 206 days [184-234] after the second dose), and IV.) uninfected and short-term three-dose vaccinated individuals (Vx3, n=5, a median of 52 days [42-54] after the third dose or boost). Whereas our data showed that S-specific T cells can be detected in the lung of mRNA-vaccinated individuals up to 8 months after immunization, lung responses in vaccinated patients presented less frequently a T_RM_ phenotype and polyfunctional T_RM_ expressing IFNγ and CD107a were essentially absent compared to convalescent patients.

## Results

### Cohort characteristics

Paired cross-sectional peripheral blood and healthy tissue areas obtained from patients undergoing lung resection for differing oncologic reasons were studied. Patient characteristics are summarized in Supplemental Table 1. In order to confirm the SARS-CoV-2 status of each patient, we analyzed total immunoglobulin (Ig) or IgG levels against N and Spike (S) proteins respectively, which discriminated Ctrl (negative for N and S), Inf patients (positive for N and S) and vaccinated groups (negative for N and positive for S; Supplemental Table 1). Furthermore, the viral neutralization titer was determined against the SARS-CoV-2 Omicron variant using a pseudovirus neutralization assay and, as expected ^10, 11^, a positive correlation between neutralization and S-IgG titers was detected (Spearman r = 0.72, *P* = 0.0016; Supplemental Figure 1A). In addition to the absence of neutralization of the Omicron variant in plasma of the Ctrl group, 2 out of 7 patients (28%) in the Inf group and from 1 out of 6 patients (17%) in the Vx2 group failed to neutralize the virus, whereas all patients in the Vx3 group were able to neutralize this variant (Supplemental Table 1). The fact that we mostly studied elderly patients could certainly determine the overall response and, indeed there was a negative correlation between older age and neutralizing capacity for the Inf group (Spearman r = - 0.88, *P* = 0.01; Supplemental Figure 1B) and the same trend was observed for the Vx2 group (Spearman r = - 0.72, *P* = 0.10; Supplemental Figure 1C). This relationship was less evident between age and S-IgG titers (Supplemental Figure 1D, E), yet examples in larger cohorts exist ^10^. Instead, S-IgG titers from all groups negatively correlated with sample timing (Spearman r = -0.61, *P* = 0.010; Supplemental Figure 1F), a correlation that was also observed for total Ig against N in the Inf group (Spearman r = -0.88, *P* = 0.009; Supplemental Figure 1G), which agrees with antibody titers decay ^10, 11, 12^.

### Recent mRNA booster vaccination elicits S-specific CD4^+^ T cells similar to convalescent infection

To address cellular immune responses, we stimulated fresh peripheral blood mononuclear cells (PBMC) and lung-derived cellular suspensions with overlapping Membrane (M), N and S peptide pools and determined the intracellular expression of IFNγ, interleukin (IL)-4, and IL-10, along with the degranulation marker CD107a in CD4^+^ and CD8^+^ T cells (Supplemental Figure 2A), as previously described ^8^. We found detectable circulating IFNγ-secreting Ag-specific CD4^+^ T cells responding to all proteins in the blood of Inf patients, which was significantly higher compared to the Ctrl, Vx2, and Vx3 groups for M and N peptides (Figure 1A, B). In contrast, for S peptides, the Inf group only showed higher frequencies of IFNγ^+^ CD4^+^ T cells compared to the Ctrl group, indicating an increase induced by vaccination in the blood of Vx2 and Vx3 groups. However, only two Vx2 patients showed detectable frequencies of S-specific CD4+ T cells in blood, while recently boosted Vx3 patients displayed an overall increase reaching statistical significance compared to the Ctrl group (Figure 1B). In contrast to CD4^+^ T cells, the frequencies of IFNγ^+^ CD8^+^ T cells detected were minimal for each of the groups against any of the proteins, including for the Vx groups against S peptides (Figure 1B). Expression of IL-4, IL-10, and CD107a by T cells showed, in general, high variability, limiting the detection of differences (Supplemental Figure 3). Nonetheless, S-specific degranulating CD107a^+^ CD8^+^ T cells were overall more frequent in the Vx2 compared to the Ctrl group (*P* = 0.046; Supplemental Figure 3). Together, these data indicate that M, N, and S-peptide specific IFNγ^+^ CD4^+^ T cell responses can be readily detected in blood months after resolving natural SARS-CoV-2 infection and that these responses require a recent mRNA vaccine booster-dose against SARS-CoV-2 to elicit similar frequencies against the S protein in vaccinated individuals.

**Figure 1.**
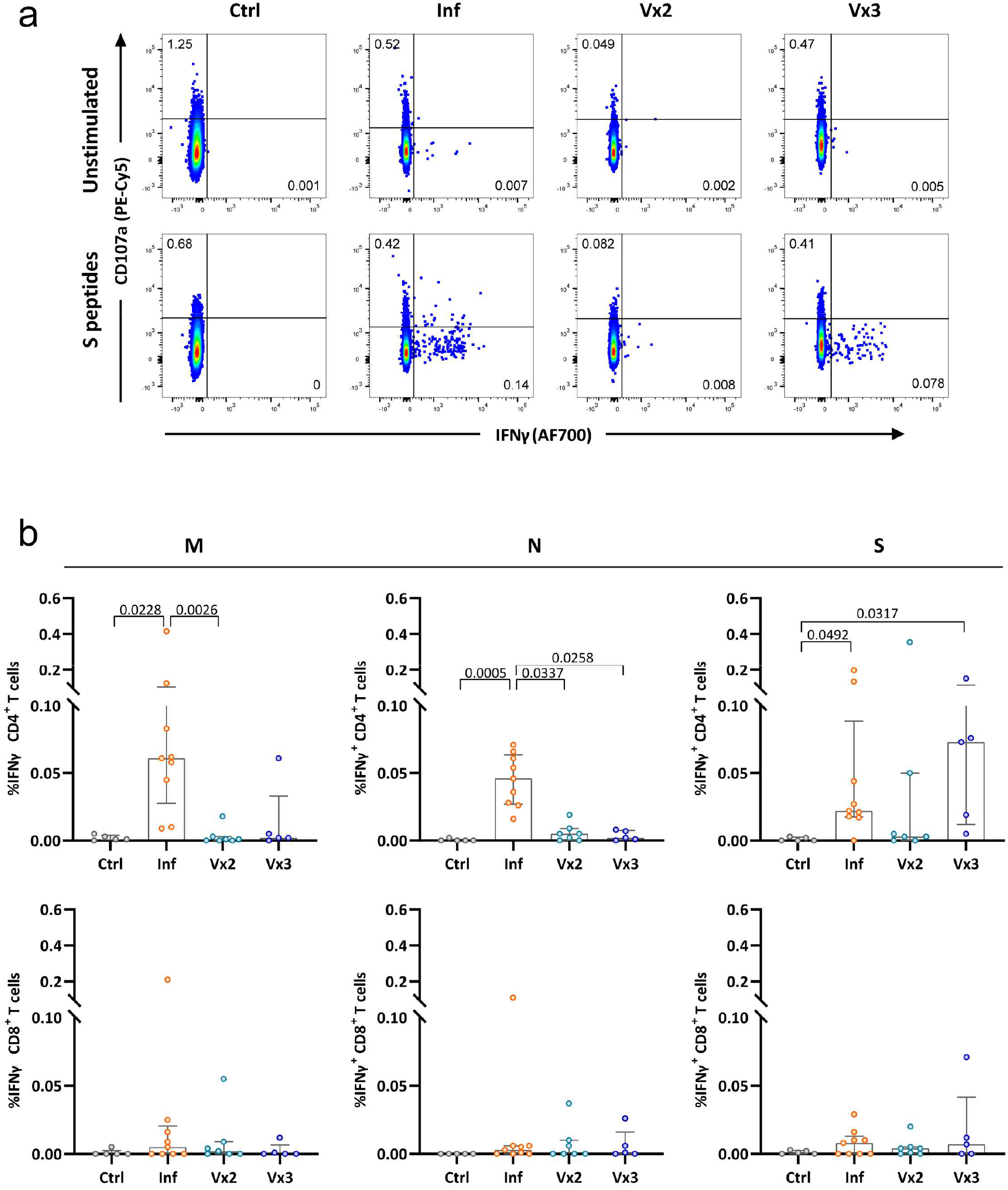
SARS-CoV-2-specific T-cell responses in peripheral blood from convalescent and vaccinated patients. (**A**) Representative flow-cytometry plots showing CD4^+^ T cells expressing CD107a and IFNγ after exposure of whole PBMCs to S-peptide pools or left unstimulated for each of the four groups included in this study (complete gating strategy is shown in Supplemental Figure 2A). (**B**) Comparison of the net frequency (background subtracted) of IFNγ^+^ cells within CD4^+^ (upper) and CD8^+^ (lower) T-cell subsets after stimulation of PBMCs with any of the three viral peptide pools (membrane (M), nucleocapsid (N) and spike (S) peptides). Data are shown as median ± IQR, where each dot represents an individual patient for each group (Ctrl, control, n=5; Inf, convalescent infected, n=9; Vx2, vaccine 2 doses, n=7 and Vx3, vaccine 3 doses, n=5). Statistical significance was determined by Kruskal-Wallis test (with Dunn’s post-test).

### mRNA vaccination induces S-specific CD4^+^ T cells in the lung

As reported previously ^8^, we here found that mild or severe natural infection with SARS-CoV-2 induced robust IFNγ^+^ CD4^+^ T cells in the lung against M, N, and S peptides, detectable for up to 12 months after infection (Figure 2A, B). Interestingly, whereas M and N-specific IFNγ^+^ CD4^+^ T-cell frequencies were significantly higher in the Inf group compared to Ctrl or Vx groups, these differences were not observed for S-specific responses (Figure 2A, B). Vx2 and Vx3 groups showed presence of S-specific IFNγ^+^ CD4^+^ T cells in the lung in most patients and its frequency was comparable to levels detected in Inf patients, although statistical significance was no reached compared to the Ctrl group (Figure 2B). In contrast to CD4^+^ T cells, CD8^+^ T cells producing IFNγ after stimulation with M, N, or S peptides was variable within each group and did not result in significant differences between the groups, indicating that natural infection nor vaccination elicit a robust IFNγ positive CD8^+^ T cell response in the human lung (Figure 2B). Furthermore, induction of lung anti-SARS-CoV-2-specific T-cell responses involving expression of IL-4, IL-10, and CD107a did not differ between groups (Supplemental Figure 4A). Of note, in this tissue compartment, we detected negative correlations between patient’s age within the Inf group and the frequency of S-specific degranulating CD4^+^ and CD8^+^ T cells (Spearman r = - 0.76, *P* = 0.024 and Spearman r = - 0.77, *P* = 0.020 respectively, Supplemental Figure 4B).

**Figure 2.**
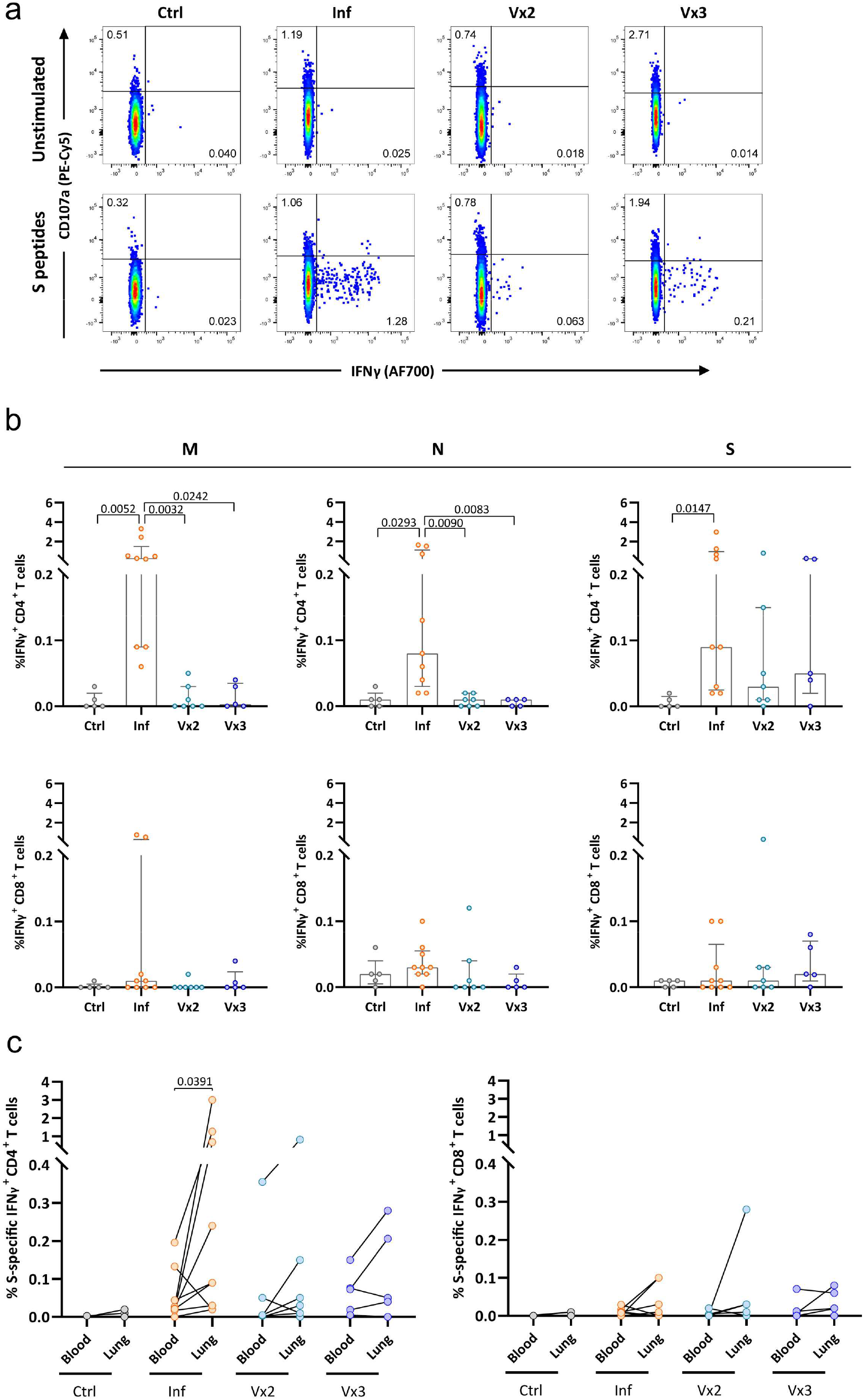
SARS-CoV-2-specific lung T-cell responses from convalescent and vaccinated patients and comparison between tissue compartments. (**A**) Representative flow-cytometry plots showing CD4^+^ T cells expressing CD107a and IFNγ after exposure of single-cell suspensions of lung tissue to S-peptide pools or left unstimulated for each of the four groups included in this study (complete gating strategy is shown in Supplemental Figure 2B). (**B**) Comparison of the net frequency (background subtracted) of IFNγ^+^ cells within CD4^+^ (upper) and CD8^+^ (lower) T-cell subsets after exposure of lung single-cell suspensions to any of the three viral peptide pools (membrane (M), nucleocapsid (N) and spike (S) peptides). (**C**) Comparison of the net frequency of IFNγ^+^ cells within CD4^+^ (left) and CD8^+^ (right) T-cell subsets in paired blood and lung samples of each group after exposure to S-peptide pools. Data in bar graphs are shown as median ± IQR, where each dot represents an individual patient for each group (Ctrl, control, n=5; Inf, convalescent infected, n=9; Vx2, vaccine 2 doses, n=7 and Vx3, vaccine 3 doses, n=5). Statistical significance was determined by (**B**) Kruskal-Wallis test (with Dunn’s post-test) or (**C**) Friedmann test (with Dunn’s post-test).

When we compared the magnitude of S-specific T cells in paired blood and lung samples, we found increased frequencies of IFNγ^+^ CD4^+^ T cells in the lungs of patients from the Inf group compared to blood (*P* =0.039, Figure 2C). The same trend was observed for the Vx individuals, which was close to significant if both groups were pooled (*P* = 0.054), although this increase was more variable as only 7 out of 12 Vx patients showed an increase, in contrast to 8 out of 9 Inf patients. In contrast, the CD8^+^ T-cell compartment did not show clear differences between these two compartments (Figure 2C), neither any of the T-cell subsets for any other function, which were highly variable (Supplemental Figure 5). Together our data indicates that S-specific CD4^+^ T-cell responses are detectable in the lung of uninfected vaccinated patients, suggesting that mRNA vaccination against SARS-CoV-2 may potentially elicit tissue-localized protective T-cell responses already after the second mRNA vaccine dose.

### Limited induction of tissue resident memory T cells by mRNA vaccination

Presence of T_RM_ cells may provide a better correlate of protection from disease in SARS-CoV-2 infected individuals and are characterized by expression of CD69 and/or CD103 ^8 9^. Moreover, T_RM_ cells require downregulation of the transcription factor T-bet for expression of CD103 and their formation and survival at tissue sites ^13^. In order to assess if S-specific T cell responses detected in Vx patients had indeed a T_RM_ phenotype, we analyzed expression of CD69 and CD103 by lung SARS-CoV-2-specific CD4^+^ and CD8^+^T cells, which we classified as: CD69^-^ (non-T_RM_), CD69^+^ (T_RM_) and a subset within CD69^+^ cells expressing CD103^+^ (T_RM_ CD103^+^) (Figure 3A, Supplemental Figure 2B for gating strategy). Additionally, we assessed expression of T-bet to assure that CD69 in the lung was associated to a T_RM_ phenotype and not to activation of CD69^-^ T cells or the product of residual blood in the lung. Both CD69^+^ T_RM_ and CD103^+^ T_RM_ subsets did not show T-bet expression across all patient groups, whereas a fraction of CD69^-^ non-T_RM_ cells presented T-bet expression (Supplemental Figure 2B), suggesting the association to tissue residency when absent ^8^. S-specific CD4^+^ T cells from the Inf group showed higher frequencies of IFNγ^+^ cells within the CD69^+^ and CD103^+^ T_RM_ phenotypes (Figure 3A, B), with statistical significance reached for the overall CD69^+^ T_RM_ fraction compared to the non-T_RM_ fraction. Furthermore, while no significant differences were detected for CD103^+^ T_RM_ cells against S-peptides in any of the groups, a trend was observed for CD4^+^ T-cell responses to M peptides and statistical significance was reached for CD8^+^ T cells against N peptides compared to the non-T_RM_ fraction in the Inf group (Supplemental Figure 6A, B). Of note, a negative correlation was observed between IFNγ-secreting S-specific CD8^+^ CD103^+^ T_RM_ cells and sample timing (Spearman r = -0.82, *P* = 0.019 Supplemental Figure 6C). In addition, some patients in the Vx2 and Vx3 groups showed modest presence of S-specific T_RM_ with or without CD103 expression in their lungs (Figure 3A, B). However, this response was highly heterogeneous and not statistically significant. These findings indicate that mRNA vaccination against SARS-CoV-2 can induce S-specific T_RM_ in some, but not all individuals and may also last long term after the second vaccination.

**Figure 3.**
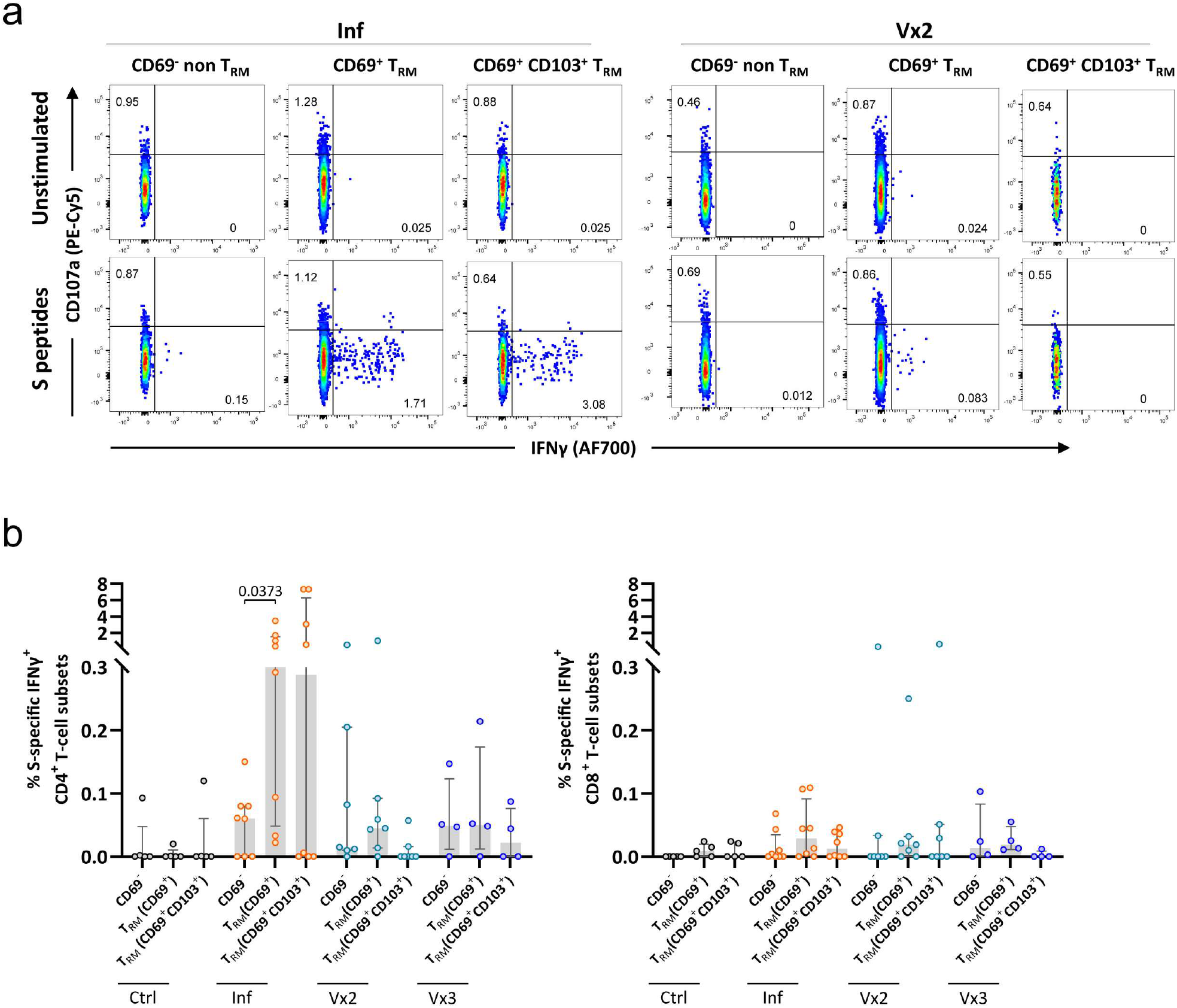
Frequency of Spike-specific T_RM_ cells in the lung. (**A**) Representative flow-cytometry plots showing three subsets of CD4^+^ T cells present in the lung: CD69^-^ non-T_RM_, CD69^+^ T_RM_, and CD69^+^CD103^+^ T_RM_ cells expressing CD107a and IFNγ after exposure of single-cell suspensions of lung tissue to S-peptide pools or left unstimulated for an Inf and a Vx2 patients. (**B**) Comparison of the net frequency of S-specific IFNγ^+^ cells within the three (non-) T_RM_ cell subsets present in the lung for each group. Data in bar graphs are shown as median ± IQR, where each dot represents an individual patient for each group (Ctrl, control, n=5; Inf, convalescent infected, n=8; Vx2, vaccine 2 doses, n=7 and Vx3, vaccine 3 doses, n=4). Statistical significance was determined by Friedmann test (with Dunn’s post-test) for the difference between the cellular subsets within each patient group.

### Overall functional T-cell response of lung and blood compartments

To better gain insight into the overall S-specific response by each group, including all functions and considering lung-T_RM_ phenotypes, we represented S-specific CD4^+^ and CD8^+^ T cell subsets as donut charts displaying the mean frequency of responses including all individuals (responders and non-responders, Figure 4). This way, a dominance of IFNγ-secreting CD4^+^ T cells was particularly associated to the two T_RM_ phenotypes in the Inf and, to a lesser extent, in the Vx2 patients (Figure 4A). Further, S-specific responses within non-T_RM_ and blood CD4^+^ T cells were functionally similar and in general dominated by IFNγ and IL-4 secretion (Figure 4A). In contrast, degranulation characterized the majority of lung S-specific CD8^+^ T cells from Inf individuals (Figure 4B), which correlated negatively with older age for the T_RM_ fractions (Spearman r = - 0.88, *P* = 0.006 for both CD103 positive and negative, Supplemental Figure 6D). Degranulation was also the major function in blood from the two Vx groups (Figure 4B). Last, in general, CD8^+^ T-cell responses considering all functions were of higher magnitude in long-term Vx2 individuals, reaching statistical significance for blood responses in comparison to the Ctrl group, as shown in the adjoin graph on the right (Figure 4B).

**Figure 4.**
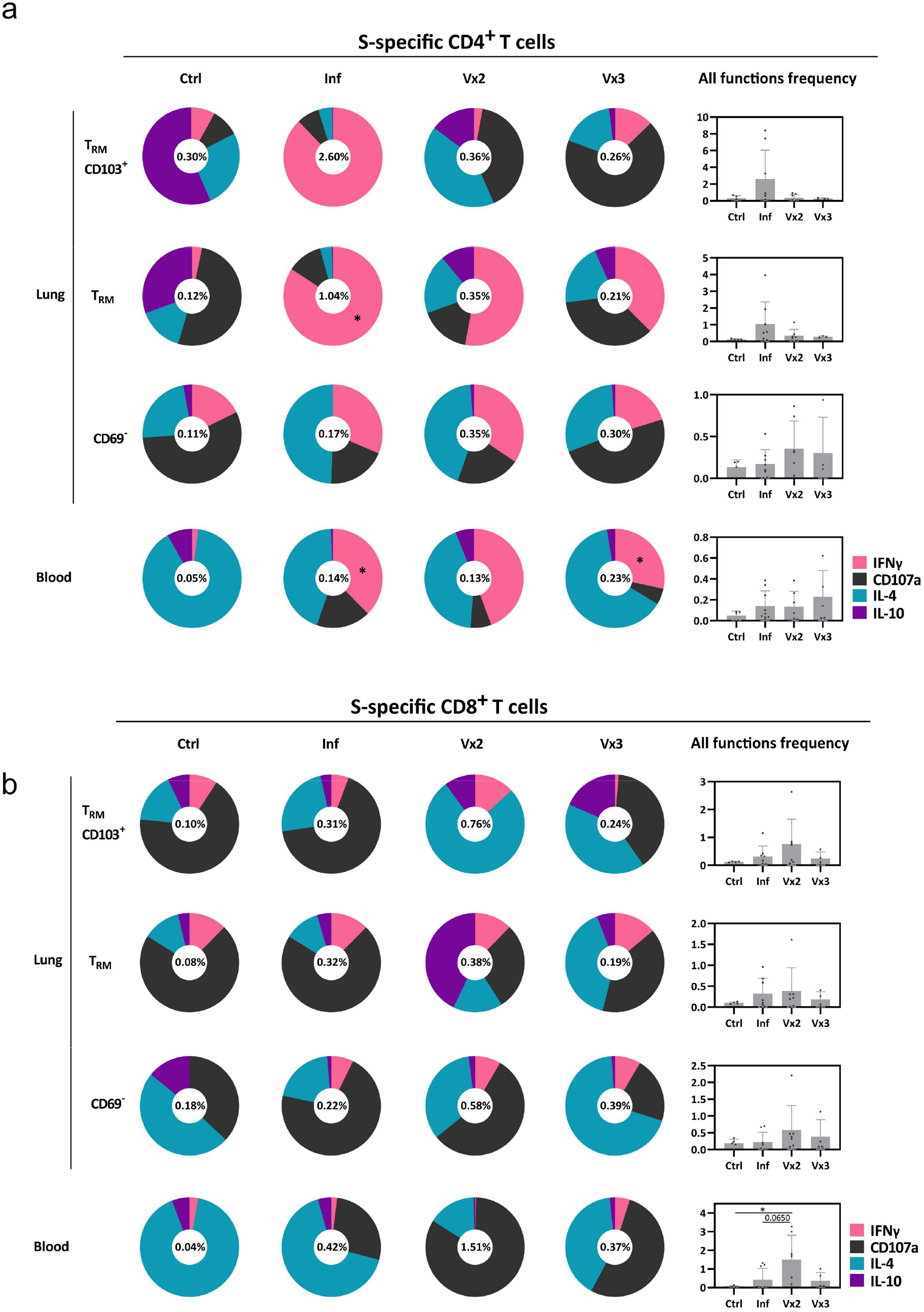
Overall functional T-cell response of lung and blood compartments. (**A, B**) Donut charts displaying the net contribution of each functional marker (IFNγ, CD107a, IL-4, and IL-10) to the overall S-specific CD4^+^ (**A**) and CD8^+^ (**B**) T-cell response within the lung resident and non-resident T-cell subsets and in peripheral blood for each of the individual patient groups. Data represent the mean value of the net frequency of each function within the patient group, including both responders and non-responders. The frequency shown inside each donut chart represents the accumulated mean response of all functions. Bar charts on the right show the mean of the total frequency considering all functions per group (mean ± SD). Statistical significance was determined by Kruskal-Wallis test (with Dunn’s post-test) for the difference between each group. * *P* < 0.05.

### Polyfunctional S-specific IFNγ^+^CD107a^+^ CD4^+^ T cells are absent in vaccinated patients

We previously detected a low but consistent polyfunctional IFNγ^+^CD107a^+^ T-cell response mostly associated to the T_RM_ fraction in convalescent infected patients ^8^. We therefore investigated whether mRNA vaccination against SARS-CoV-2 would also induce such S-specific polyfunctional responses in both compartments (Figure 5A, B). Indeed, increased frequencies of polyfunctional IFNγ^+^CD107a^+^ CD4^+^ T cells were detected in blood from the Inf group against N peptides compared to the Ctrl group, but not against M- and S-peptides. Interestingly, a trend towards higher frequencies of S-specific polyfunctional CD4^+^ T cells was observed for the Vx3 group (Figure 5A). Likewise, circulating polyfunctional S-specific CD8^+^ T cells were enhanced in Vx2 individuals compared to Ctrl group (Figure 5A). In fact, if Vx groups were pooled to increase sample size, then both CD4^+^ and CD8^+^ T cells reached significance compared to Ctrl samples (*P* = 0.037 for CD4^+^ and *P* = 0.024 for CD8^+^). In addition, the frequency of polyfunctional IFNγ^+^CD107a^+^ cells present in total CD4^+^ and CD8^+^ T cells in the lung were only consistently increased in the Inf group against N peptides compared to the Ctrl and Vx3 groups (Figure 5B). Nevertheless, while a high degree of variability was observed among vaccinated patients, polyfunctional S-specific T cells were detected in some individuals (Figure 5B). Strikingly, S-specific CD4^+^ polyfunctional CD103^+^ T_RM_ cells were virtually absent in the Vx2 and Vx3 groups, while being frequently present in the lungs of patients from the Inf group (Figure 6). Furthermore, the frequency of S-specific polyfunctional CD4^+^ T cells in the CD69^+^ T_RM_ cells was higher in the Inf group compared to the Ctrl and Vx2 groups (Figure 6). Together, these data indicate that both short- and long-term vaccination do not induce S-specific IFNγ^+^CD107a^+^ CD103^+^ T_RM_ cells in the lung, which may contribute to antiviral activity.

**Figure 5.**
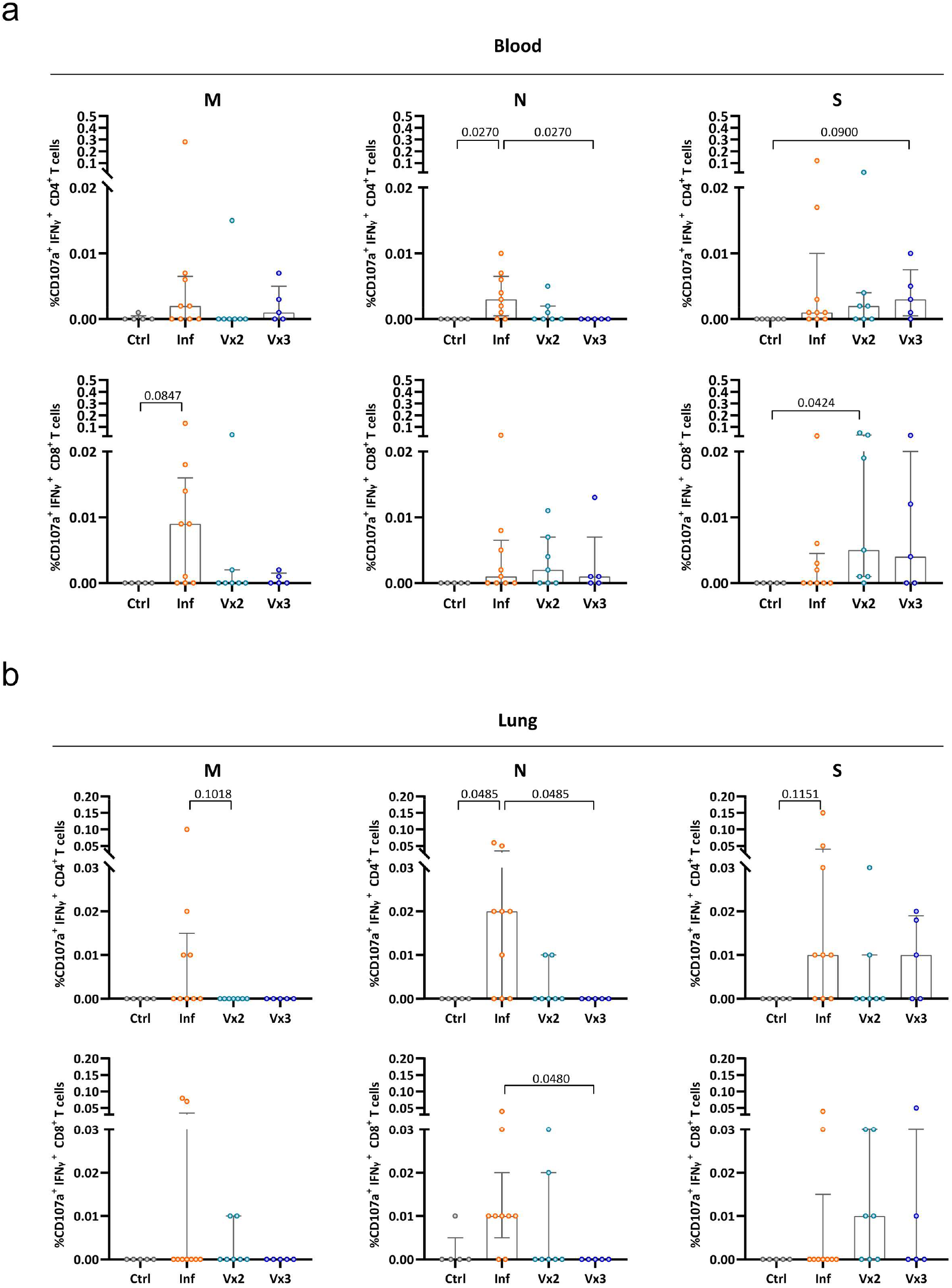
Polyfunctional CD4^+^ and CD8^+^ T-cell responses in blood and lung of convalescent and vaccinated patients. (**A, B**) Comparison of the net frequency of polyfunctional CD107a^+^ IFNγ^+^ cells within CD4^+^ and CD8^+^ T-cell subsets for each of the four groups after exposure of PBMCs (**A**) or single-cell suspensions of lung tissue (**B**) to any of the three viral peptide pools (membrane (M), nucleocapsid (N) and spike (S) peptides). Data in bar graphs are shown as median ± IQR, where each dot represents an individual patient for each group (Ctrl, control, n=5; Inf, convalescent infected, n=9; Vx2, vaccine 2 doses, n=7 and Vx3, vaccine 3 doses, n=5). Statistical significance was determined by was determined using Kruskal-Wallis test (with Dunn’s post-test).

**Figure 6.**
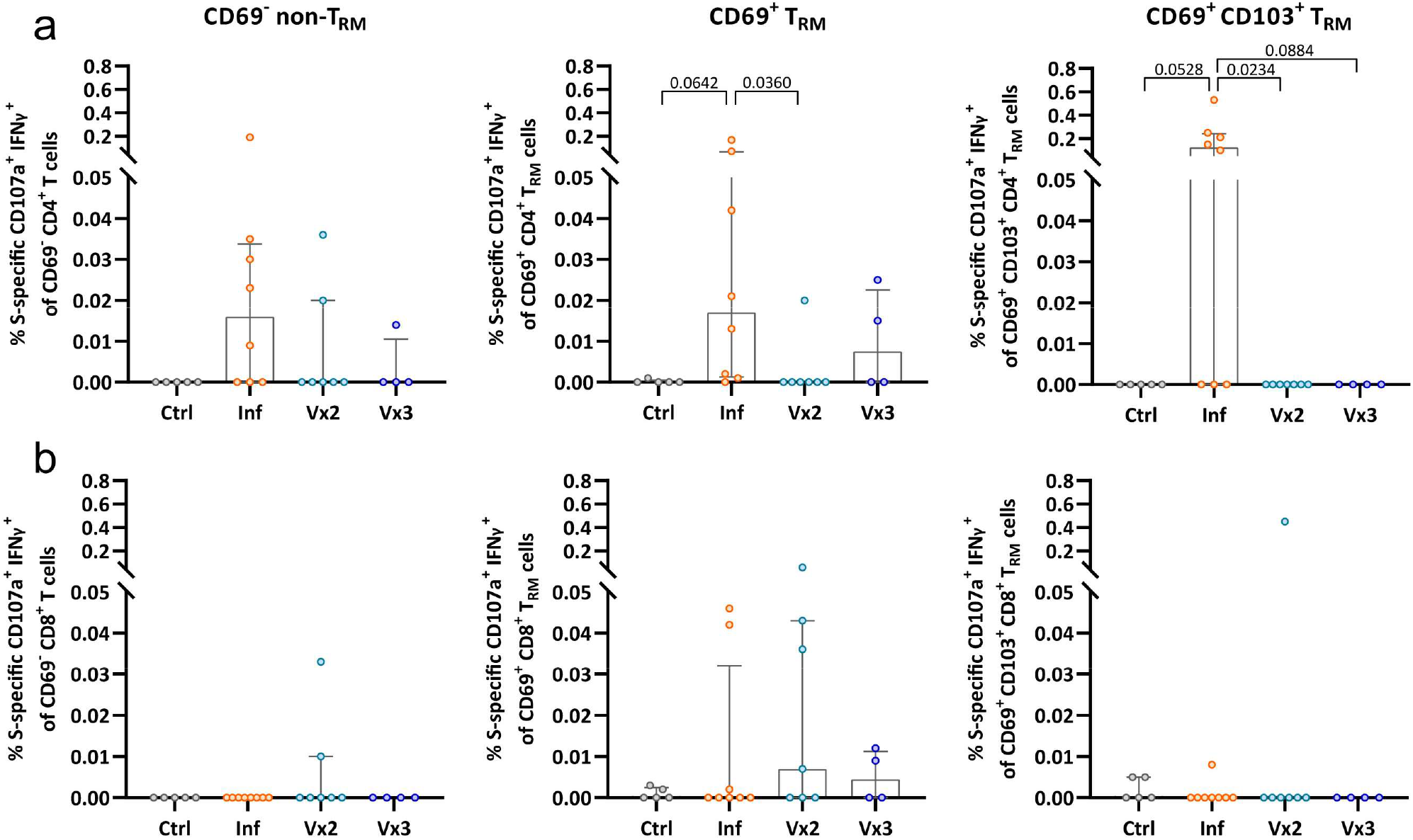
Frequency of polyfunctional T-cell responses against spike with a tissue-resident phenotype in the lung. Comparison of the net frequency of S-specific polyfunctional CD107a^+^ IFNγ^+^ cells within lung CD4^+^ (upper) and CD8^+^ (lower) (non-) tissue-resident cell subsets (CD69-non-T_RM_, CD69^+^ T_RM_, and CD69^+^CD103^+^ T_RM_ cells) for each of the four patient groups after exposure of single-cell lung suspensions to S-peptide pools. Data in bar graphs are shown as median ± IQR, where each dot represents an individual patient for each group (Ctrl, control, n=5; Inf, convalescent infected, n=8; Vx2, vaccine 2 doses, n=7 and Vx3, vaccine 3 doses, n=4). Statistical significance was determined by Kruskal-Wallis test (with Dunn’s post-test) for the difference between the groups.

### Anti-SARS-CoV-2 T cell response dynamics in two cases of interest

Last, considering the uniqueness of analyzing immune responses in paired blood and lung parenchyma samples and recent studies detailing changes in T cell responses in infected individuals already vaccinated and vice versa ^14^, we highlight two patients that were discarded due to not fitting inclusion criteria, yet bring interesting data to the study. HL174 was a patient in their fifties who received the third mRNA-1273 vaccine boost and, five days after, tested positive by PCR. We analyzed paired tissue samples 30 days after the boost/infection event (Supplemental Figure 7A). This patient had a neutralization titer of 1740 IU/mL against omicron, and had detectable IgG and Ig titers against S and N proteins (>800 AU/mL and 1.23 index, respectively). When comparing T-cell responses from blood and lung tissue, a much higher IFNγ-response was observed in the lung, in particular against the N protein, which already contained responding cells with a T_RM_ phenotype (Supplemental Figure 7B, C). In contrast, in blood, degranulation was enhanced mostly against S but also M protein and some proportion of IL-10 secretion was detected against all proteins (Supplemental Figure 7B).

On the other hand, patient HL162, who was in their early seventies, was first infected presenting a mild COVID-19 and, several months after, received three doses of the mRNA-1273 vaccine. In this case, we obtained samples 3.7 months after infection and another one 1.3 months after the third dose (due to a second intervention for a lung carcinoma), which corresponded to a year after initial infection, as shown (Supplemental Figure 8A). Neither of these two time points showed neutralization titers against omicron and the titers of IgG, instead of increasing after triple vaccination, decreased from 156 to 0 index for the N protein and from 306.54 to 13.85 AU/mL for S protein. The comparison of the tissue compartments after infection and after triple vaccination evidenced a concomitant strong decrease in T-cell responses in blood and tissue (Supplemental Figure 8B, C, and Supplemental Figure 9A, B). However, IFNγ-secreting SARS-CoV-2 T cells against M and N proteins in the lung were better preserved from the original infection one year later than were responses against the S protein enhanced due to vaccination (Supplemental Figure 8B, C and Supplemental Figure 9A, B). Thus, while the lower respiratory tract compartment more faithfully represented T_RM_ responses established already during the infection event one year earlier, responses in blood mostly vanished.

### Discussion

Comprehensive studies comparing the magnitude and duration of the T cell responses indicate similar magnitude after dual vaccination and after natural SARS-CoV-2 infection ^4, 11, 14^. However, these results may not hold if we consider that the magnitude, the functional profile and even the duration of these responses in blood may not faithfully reflect responses in the respiratory tract ^6, 8, 9, 15^. In fact, the individual comparison between these two compartments among the S-responding T cells from the different groups showed higher magnitude in the lung than in the blood, but also a different profile. A key difference, and the main driver of our study, was the establishment of long-term protection potentially mediated by T_RM_ after vaccination, since longevity of SARS-CoV-2 T cell responses remains a critical question ^6^. In principle, T_RM_ are established by mucosal infection since Ag together with local signals promote the recruitment and establishment of this memory response. In this sense, intramuscular vaccination with an adenovector vaccine in mice did not induce SARS-CoV-2-specific T_RM_ in their lungs ^16^. Thus, to induce potent resident immunity, vaccine strategies may need to either use live-attenuated Ag or employ mucosal routes. Consequently, the absence of vaccine induced S-specific T_RM_ could be expected in infection-naïve individuals. Still, recent data shows that a secretory IgA response was induced in ∼30% of participants after two doses of a SARS-CoV-2 mRNA vaccine which, in addition, may play an important role in protection against infection ^15^. While we detected S-specific IFNγ^+^ CD4^+^ T cell responses in the lung of vaccinated individuals, the proportion of these cells in the T_RM_ phenotype was modest, in particular if considering CD103 expression. Further, the presence of polyfunctional IFNγ^+^CD107a^+^CD4^+^CD103^+^T_RM_ appeared to be restricted to the lungs of convalescent-infected patients only, while absent in the lungs of vaccinated patients.

Another difference in the comparison of the cellular immunity between SARS-CoV-2-infected convalescent and uninfected-vaccinated individuals is the broader and potentially stronger response induced by symptomatic infection. This is partially manifested by the fact that the overall magnitude of responses against M and N peptides are frequently higher than S peptides ^8, 17, 18, 19^. Of note, disease severity may impact both the magnitude and function of the T cell response against the different proteins ^8, 20, 21^. In addition, among other factors, age also influences the magnitude and duration of immune responses in distinct tissue compartments, even the establishment of T_RM_ ^22^, a factor that influenced the frequency of degranulation in the lung of our Inf patients, including within the T_RM_ fraction. Yet advanced age will also limit the immune response to vaccination ^23^. On the other hand, we have observed that different proteins induce different functional profiles during acute infection, which may influence disease control ^8^. S-specific immune responses may better support B cell and antibody generation via follicular helper T cells, which are instrumental to limiting infection ^4, 8^. Instead, responses against the N protein seem to more consistently induce polyfunctional antiviral T cells and these responses may be more conserved among other coronaviruses ^5, 8, 24, 25^. Indeed, preexisting SARS-CoV-2 specific T cell responses have been found in the blood of unexposed individuals ^18, 26^. In this sense, in our study we detected a low level of preexisting T cell responses in our control group mostly located in the lung, which may be in agreement with a recent report in tonsillar tissue ^27^. Thus, another conclusion would be highlighting the interest of including other proteins beyond the spike such as N sequences, which has been suggested before ^5, 8, 19, 25, 28^. Last, in terms of duration, our study lacks longitudinal data to assess the dynamics in the different compartments, yet it is assumed that T_RM_ phenotypes will contribute to long-term persistence ^6, 8, 29^. In fact, the only patient for which we had longitudinal sampling after infection and after the third vaccine boost (Supplemental Figure 8) demonstrated that even if vaccination fails to induce a systemic antibody response, a low frequency SARS-CoV-2 T cell response directed to proteins from the original infection remains exclusively detectable in the lung as T_RM_ one year after.

The overall CD8^+^ T cell response was enhanced in some but not all vaccinated patients and was in general low and dominated by degranulation. In fact, lung S-specific CD8^+^ T_RM_ presented similar overall frequencies in vaccinated individuals when considering all functions. Of note, low percentages of SARS-CoV-2 specific CD8^+^ T cells may be due to the use of 15-mer peptides, which are less optimal than 9/10-mer peptides for HLA class I binding ^3^, although this is debatable ^21^. Other methods, such as the use of activation induced markers and tetramers loaded with SARS-CoV-2 immunodominant peptides ^27, 30^ may give additional insight into the CD8^+^ T cell response against SARS-CoV-2. Thus, considering the putative protective role of CD8^+^ T cells observed in animal models ^29^, further exploration of the CD8^+^ T cell response after mRNA vaccination is warranted. our results for these vaccinated individuals are certainly encouraging.

We acknowledge that our study has several limitations. The small sample size for the different groups warrants further investigation with ideally larger cohorts. Finding patients that meet our inclusion criteria was however challenging and currently, patients meeting the criteria are no longer available. In addition, we addressed T cell immune responses in older and mostly oncologic patients. In general, T cell responses of cancer patients after vaccination against SARS-CoV-2 may be impaired ^31^, which may overall underestimate immune responses in all groups of our study. However, we still detect vaccine-induced S-specific CD4^+^ T cells in our patients, which shows that mRNA vaccination may even contribute to protection against COVID-19 in these vulnerable patients. Further, the Inf group consisted of patients recovered from mild or severe disease. Even if age and underlying conditions were similar to the other groups, disease severity may have skewed frequencies of SARS-CoV-2-specific T cells towards the higher end. Still, considering their age and condition, any of these patients with a new infection would most likely develop a more serious COVID-19 event compared to the general population. In addition, the boosted Vx3 group was sampled short term comparing to the Vx2 group, but enhancement of T cell responses would be better detected 5-10 days after boosting ^10, 11^ (which was a less likely time for scheduling surgery). Still it was enough time to suggest that there was no major enhancement of long-term durable T cell response in the lung by a third boost. Further, we did not assess the contribution of T cells targeting mutation regions to the total spike since we aimed to compare the strength and function of vaccinated and naturally infected patients (these last group obtained during the first wave). However, the overall contribution of T cell responses to mutational regions/total spike responses has been reported to be low ^14, 32^.

Overall, our results contribute to the understanding of disease protection mediated by current mRNA vaccines. While our data indicate a more robust and broader cellular response in convalescent patients, S-specific T cells can be detected in the lung of vaccinated individuals to similar overall levels 8 months after immunization, highlighting the durability of this immune arm. Further, while we detected increased levels of IFNγ^+^ T cell responses in blood after the third dose, limited benefit of boosting towards the enhancement of T cell responses in the lung was evidenced by our data. However, elderly people not responding to vaccination have been shown to benefit from a third dose ^23^ and there is an obvious benefit of boosting to provide a higher degree of antibody-mediated protection from infection in the context of high incidence of VOC ^1^. Still, if virus neutralization is unable to completely block infection, a more robust and broader T_RM_ response established in the lung of convalescent-infected individuals may have more chances of limiting disease. In this sense, polyfunctional CD107a^+^ IFNγ^+^ cells may contribute to infection clearance or even limit the occurrence of breakthrough SARS-CoV-2 infections vaccination and absence of these cells in vaccinated patients underlines the need for the development of mucosal vaccines ^33^, recently shown to be effective in inducing sterilizing immunity in mice ^34^.The inclusion of other protein fragments such as nucleocapsid peptides ^5, 8, 19, 25, 28^ in combination with mucosal routes ^35^ will likely contribute to the establishment of optimal memory T cells in future vaccine strategies.

## Methods

### Ethics statement

This study was performed in accordance with the Declaration of Helsinki and approved by the corresponding Institutional Review Board (PR(AG)212/2020) of the Vall d’Hebron University Hospital (HUVH), Barcelona, Spain. Written informed consent was provided by all patients recruited to this study.

### Subject recruitment and sample collection

Patients undergoing lung resection for various reasons at the HUVH were recruited through the Thoracic Surgery Service and invited to participate. Initially, a total of 32 patients, from whom paired blood samples and lung biopsies were collected were assayed. However, based on vaccination and/or infection status of the recruited patients, 26 (+2: HL174 and HL162) patients were finally included. Supplemental Table 1 summarizes relevant information from included patients. For all participants, whole blood was collected with EDTA anticoagulant. Plasma was collected and stored at −80 °C (except for 4 patients distributed among the different groups, as indicated in Supplemental Table 1, for which this sample was not available) and PBMCs were isolated via Ficoll–Paque separation and processed immediately for stimulation assays.

### Phenotyping and Intracellular Cytokine Staining of lung biopsies

Immediately following surgery, healthy areas from patients undergoing lung resection were collected in antibiotic-containing RPMI 1640 medium and processed as published^8^. Briefly, 8-mm^3^ dissected blocks were first enzymatically digested with 5 mg/ml collagenase IV (Gibco) and 100µg/ml of DNase I (Roche) for 30 min at 37 ºC and 400 rpm and, then, mechanically digested with a pestle. The resulting cellular suspension was first filtered through a 70µm pore size cell strainer and then filtered through a 30µm pore size cell strainer (Labclinics). After washing with PBS, cells were stimulated in a 96-well round-bottom plate for 16 to 18 hours at 37°C with 1µg/mL of SARS-CoV-2 peptides (PepTivator SARS-CoV-2 M, N or S, Miltenyi Biotec) in the presence of 3.3μL/mL α-CD28/CD49d (clones L293 and L25), 0.55μL/mL Brefeldin A, 0.385μL/mL Monensin and 5 μL/100μL anti-CD107a-PE-Cy5 (all from BD Biosciences). For each patient, a negative control, cells treated with medium, and positive control, cells incubated in the presence of 0.4nM PMA and 20μM Ionomycin, were included. Next day, cellular suspensions were stained with Live/Dead Aqua (Invitrogen) and anti-CD103 (FITC, Biolegend), anti-CD69 (PE-CF594, BD Biosciences), anti-CD40 (APC-Cy7, Biolegend), anti-CD8 (APC, BD Biosciences), anti-CD3 (BV650, BD Biosciences) and anti-CD45 (BV605, BD Biosciences) antibodies. Cells were subsequently fixed and permeabilized using the FoxP3 Fix/Perm kit (BD Biosciences) and stained with anti-IL-4 (PE-Cy7, eBioscience), anti-IL-10 (PE, BD Biosciences), anti-T-bet (BV421, Biolegend) and anti-IFNγ (AF700, Invitrogen) antibodies. After fixation with PBS 2% PFA, cells were acquired in a BD LSR Fortessa flow cytometer (Cytomics Platform, High Technology Unit, Vall d’Hebron Institut de Recerca).

### Phenotyping and Intracellular Cytokine Staining in blood

Freshly isolated PBMCs were labelled for CCR7 (PE-CF594, BD Biosciences) and CXCR3 (BV650, BD Biosciences) for 30 min at 37ºC. After washing with PBS, PBMCs were stimulated in a 96-well round-bottom plate for 16 to 18 hours at 37°C with 1µg/mL of SARS-CoV-2 peptides together with the same concentration of Brefeldin A, Monensin, α-CD28/CD49d and CD107a-PE-Cy5, as stated for the lung suspension above and published before^8^. For each patient, a negative control and a positive control were also included. After stimulation, cells were washed twice with PBS and stained with Aqua LIVE/DEAD fixable dead cell stain kit (Invitrogen). Cell surface antibody staining included anti-CD3 (Per-CP), anti-CD4 (BV605) and anti-CD56 (FITC) (all from BD Biosciences). Cells were subsequently fixed and permeabilized using the Cytofix/Cytoperm kit (BD Biosciences) and stained with anti-Caspase-3 (AF647, BD Biosciences), anti-Bcl-2 (BV421, Biolegend), anti-IL-4 (PE-Cy7, eBioscience), anti-IL-10 (PE, BD Biosciences) and anti-IFNg (AF700, Invitrogen) for 30 mins. Cells were then fixed with PBS 2% PFA and acquired in a BD LSR Fortessa flow cytometer.

### SARS-CoV-2 serology

The serological status of patients included in this study was determined in serum samples using two commercial chemiluminescence immunoassays (CLIA) targeting specific SARS-CoV-2 antibodies: (1) Elecsys Anti-SARS-CoV-2 (Roche Diagnostics, Mannheim, Germany) was performed on the Cobas 8800 system (Roche Diagnostics, Basel, Switzerland) for the determination of total antibodies (including IgG, IgM, and IgA) against nucleocapsid (N) SARS-CoV-2 protein; and (2) Liaison SARS-CoV-2 TrimericS IgG (DiaSorin, Stillwater, MN) was performed on the LIAISON XL Analyzer (DiaSorin, Saluggia, Italy) for the determination of IgG antibodies against the spike (S) glycoprotein.

### Pseudovirus neutralization assay

The spike of the Omicron SARS-CoV-2 was generated (GeneArt Gene Synthesis, ThermoFisher Scientific) from the plasmid containing the D614G mutation with a deletion of 19 amino acids, which was modified to include the mutations specific for this VOC (A67V, Δ69-70, T95I, G142D/Δ143-145, Δ211/L212I, ins214EPE, G339D, S371L, S373P, S375F, K417N, N440K, G446S, S477N, T478K, E484A, Q493R, G496S, Q498R, N501Y, Y505H, T547K, D614G, H655Y, N679K, P681H, N764K, D796Y, N856K, Q954H, N969K, L981F) (kindly provided by Drs. J. Blanco and B. Trinite). Pseudotyped viral stocks of VSV*ΔG(Luc)-S were generated following the protocol described in^36^. Briefly, 293T cells were transfected with 3µg of the omicron plasmid (pcDNA3.1 omicron). Next day, cells were infected with a VSV-G-Luc virus (MOI=1) for 2h and washed twice with warm PBS. To neutralize contaminating VSV*ΔG(Luc)-G particles cells were incubated overnight in media containing 10% of the supernatant from the I1 hybridoma (ATCC CRL-2700), containing anti-VSV-G antibodies. Next day, viral particles were harvested and titrated in VeroE6 cells by enzyme luminescence assay (Britelite plus kit; PerkinElmer). For the neutralization assays, VeroE6 cells were seeded in 96-well white, flat-bottom plates (Thermo Scientific) at 30,000 cells/well. Plasma samples were heat-inactivated and diluted four-fold towards a concentration of 1/32 of the initial sample. Diluted plasma samples were then incubated with pseudotyped virus (VSV*ΔG(Luc)-S^omicron^) with titers of approximately 1×10^6^ – 5×10^5^ RLUs/ml of luciferase activity -in a 96 well-plate flat bottom for 1 hour at 37°C, 5% CO_2_. Next, 30,000 Vero E6 cells were added to each well and incubated at 37°C, 5% CO_2_ for 20-24 hours. Then, viral entry was measured by the expression of luciferase. Cells were incubated with Britelite plus reagent (Britelite plus kit; PerkinElmer) and then transferred to an opaque black plate. Luminescence was immediately recorded by a luminescence plate reader (LUMIstar Omega). Viral neutralization was calculated as the reciprocal plasma dilution (ID50) resulting in a 50% reduction in relative light units. If no neutralization was observed, an arbitrary titer value of 16 (half of the limit of detection [LOD]) was reported.

### Statistical analyses

Flow cytometry data was analyzed using FlowJo v10.7.1 software (TreeStar). Data and statistical analyses were performed using Prism 8.0 (GraphPad Software, La Jolla, CA, USA). Data shown in bar graphs were expressed as median and Interquartile range (IQR), unless stated otherwise. Correlation analyses were performed using non-parametric Spearman rank correlation. Kruskal-Wallis rank-sum test with Dunn’s post-hoc test was used for multiple comparisons. Friedmann test with Dunn’s post-hoc test was applied for paired comparisons. A *P* value <0.05 was considered statistically significant. Antigen-specific T-cell data was calculated as the net frequency, where the individual percentage of expression for a given molecule in the control condition (vehicle) was subtracted from the corresponding SARS-CoV-2-peptide stimulated conditions.

## Data Availability

All data produced in the present work are contained in the manuscript

## Data availability

The data that support the findings of this study are available from the corresponding author upon reasonable request.

## Author contributions

Conceptualization, M.G.; Patient Recruitment and Sample Collection, J.R., V.F.; Methodology, DKJ.P., SG.K., A.G.R., J.R-C, C.M., J.E.; Investigation, DKJ.P., SG.K., A.G.R., J.E.; Formal Analysis, DKJ.P., A.G.R., M.J.B. and M.G.; Writing-Original Draft, DKJ.P. and M.G; Writing-Review & Editing, all authors; Funding Acquisition, M.G.; Supervision, M.G.

## Acknowledgements

We would like to thank all the patients who participated in the study and Drs. Julià Blanco and Benjamin Trinite for providing the plasmid encoding the omicron spike. This work was supported by grants from Fundació La Marató TV3 (201814-10 FMTV3 and 202112-30 FMTV3), from the Health department of the Government of Catalonia (DGRIS 1_5), and the Spanish AIDS network Red Temática Cooperativa de Investigación en SIDA (RD16/0025/0007). M.J.B is supported by the Miguel Servet program funded by the Spanish Health Institute Carlos III (CP17/00179). The funders had no role in study design, data collection and analysis, the decision to publish, or preparation of the manuscript.

