## Supplemental Figures 1-9 for "Limited induction of lung-resident memory T cell responses against SARS-CoV-2 by mRNA vaccination"

### SUPPLEMENTAL FIGURES AND FIGURE LEGENDS

### Supplemental Figure 1

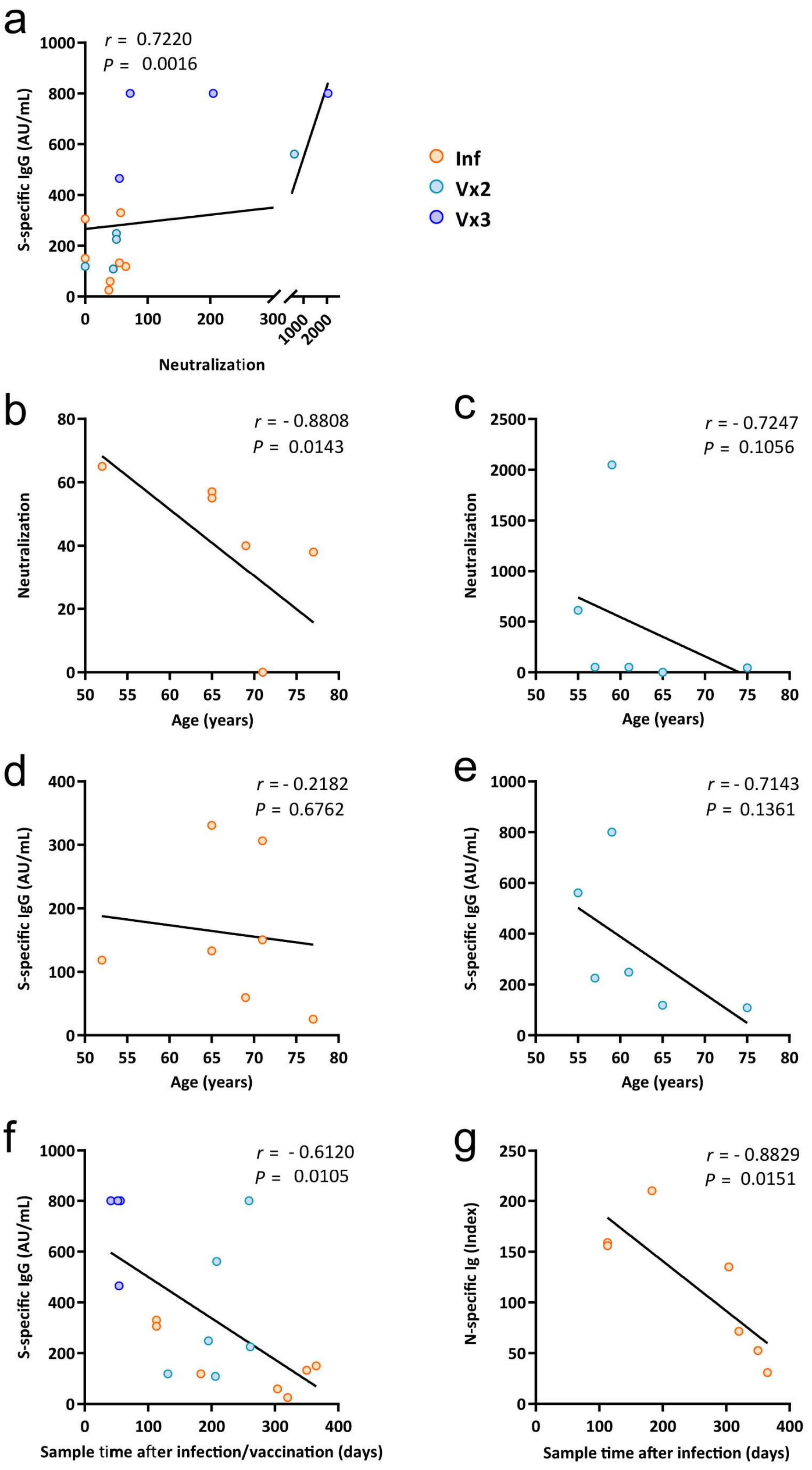

**Supplemental Figure 1. Correlation between SARS-CoV-2-specific antibodies, neutralizing capacity, age, and sampling time.** Graphs show the relationship between: (**a**) S-specific IgG antibodies (AU/mL) in plasma and SARS-CoV-2 neutralization titer for each group (Inf, convalescent infected, n=7; Vx2, vaccine 2 doses, n=6 and Vx3, vaccine 3 doses, n=4); (**b, c**) SARS-CoV-2 neutralization titer and age of Inf patients (**b**) and Vx2 patients (**c**); (**d, e**) S-specific IgG antibodies (AU/mL) and age of Inf patients (**d**) and Vx2 patients (**e**); (**f**) S-specific IgG antibodies (AU/mL) in all groups and day of sampling after infection or vaccination; and (**e**) N-specific Ig antibodies (index) in Inf patients and day of sampling after infection. Correlations (*r* and *P* values) were assessed by Spearman test.

### Supplemental Figure 2

a

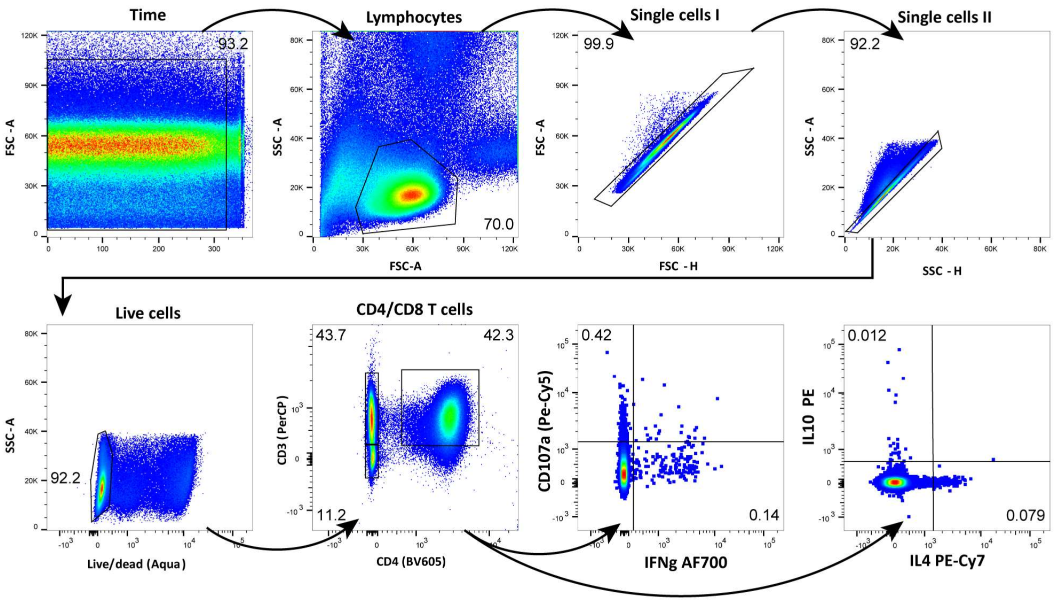


b

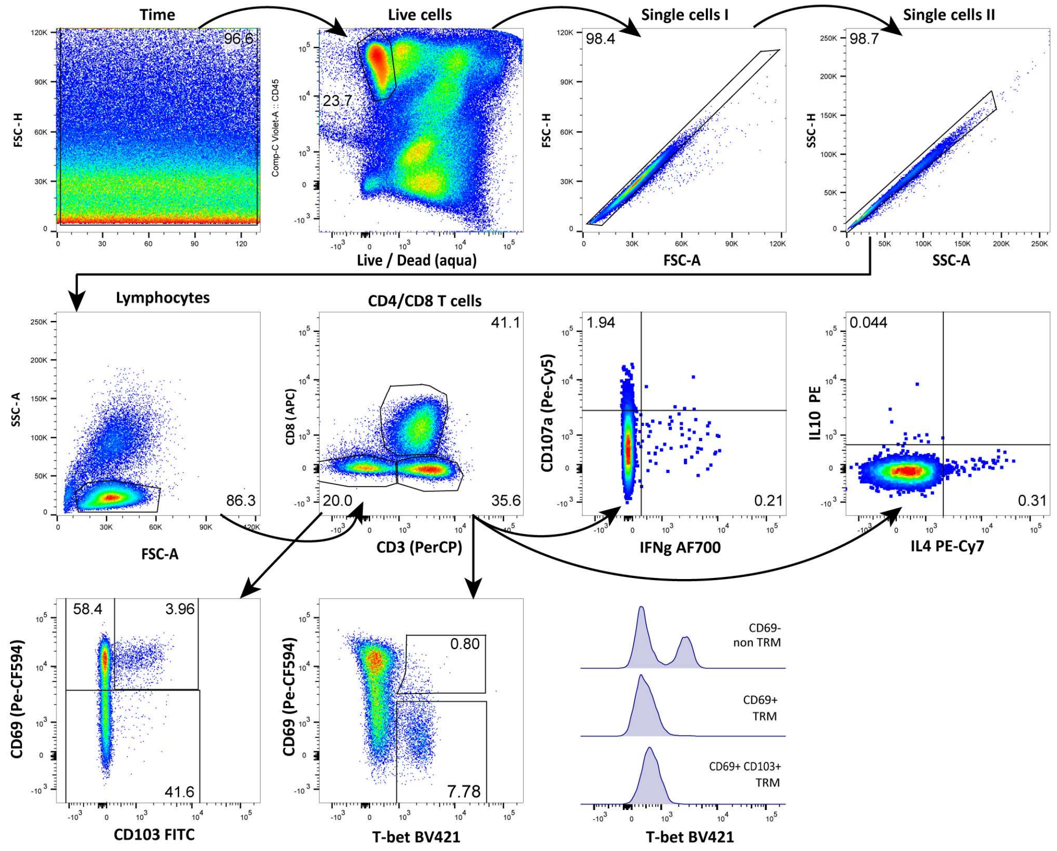

**Supplemental Figure 2. Gating strategy for the analysis of T cells present in peripheral blood and lung tissue.** (**a, b**) Representative flow-cytometry plots showing the gating strategy towards the identification of $CD4^+$ and $CD8^+$ T cells within PMBC (**a**) and lung tissue (**b**) samples. $CD4^+$ and $CD8^+$ T-cell subsets in PBMCs were identified by gating of time (to exclude disturbances in flow measurements), followed by gating of total lymphocytes, single cells, and live cells. $CD4^+$ and $CD8^+$ T-cell subsets in lung tissue were identified by gating of time, live $CD45^+$ cells, single cells, and lymphocytes.

### Supplemental Figure 3

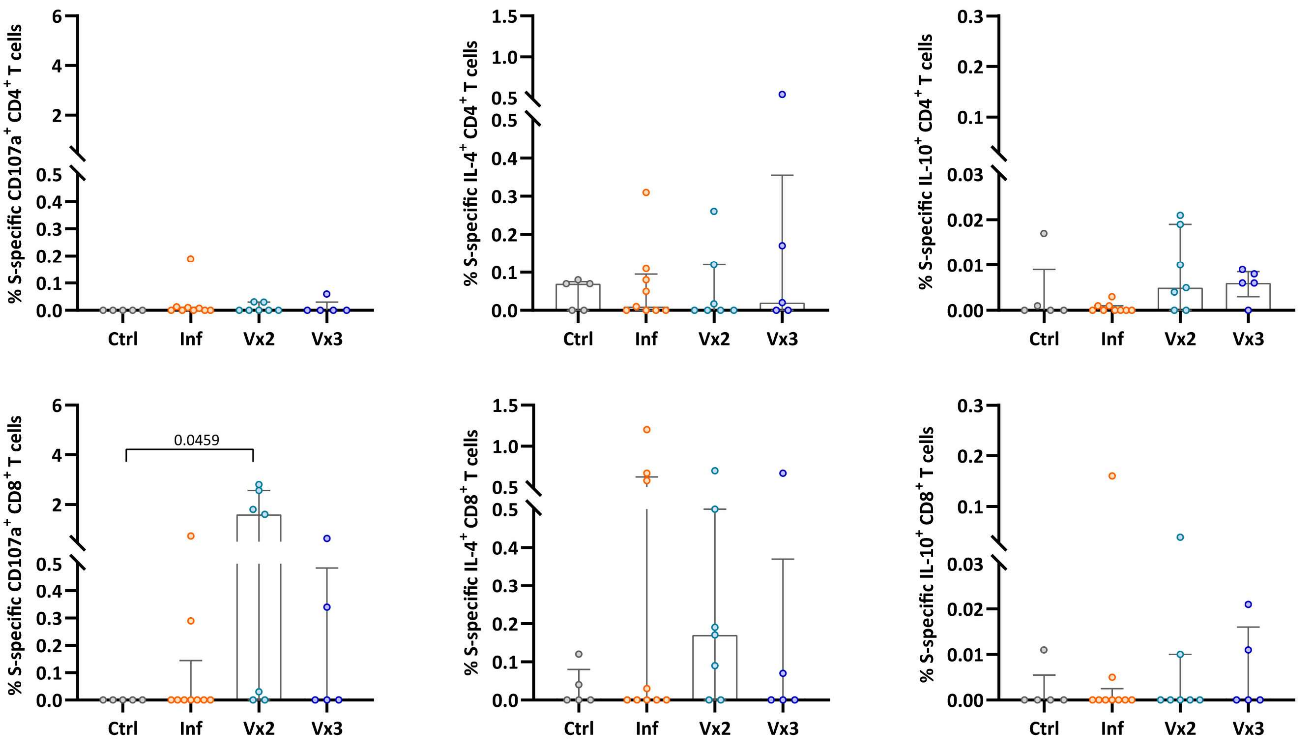


**Supplemental Figure 3. Spike-specific T-cell responses (CD107a, IL-4, IL-10) in peripheral blood from convalescent and vaccinated patients.** Comparison of the net frequency of CD107a⁺ (left), IL-4⁺ (middle), and IL-10⁺ (right) cells within CD4⁺ (upper) and CD8⁺ (lower) T-cell subsets for each of the four groups after exposure of PBMCs to S-peptide pools. Data in bar graphs are shown as median ± IQR, where each dot represents an individual patient for each group (Ctrl, control, n=5; Inf, convalescent infected, n=9; Vx2, vaccine 2 doses, n=7 and Vx3, vaccine 3 doses, n=5). Statistical significance was determined by Kruskal-Wallis test (with Dunn´s post-test).

**Supplemental Figure 4**

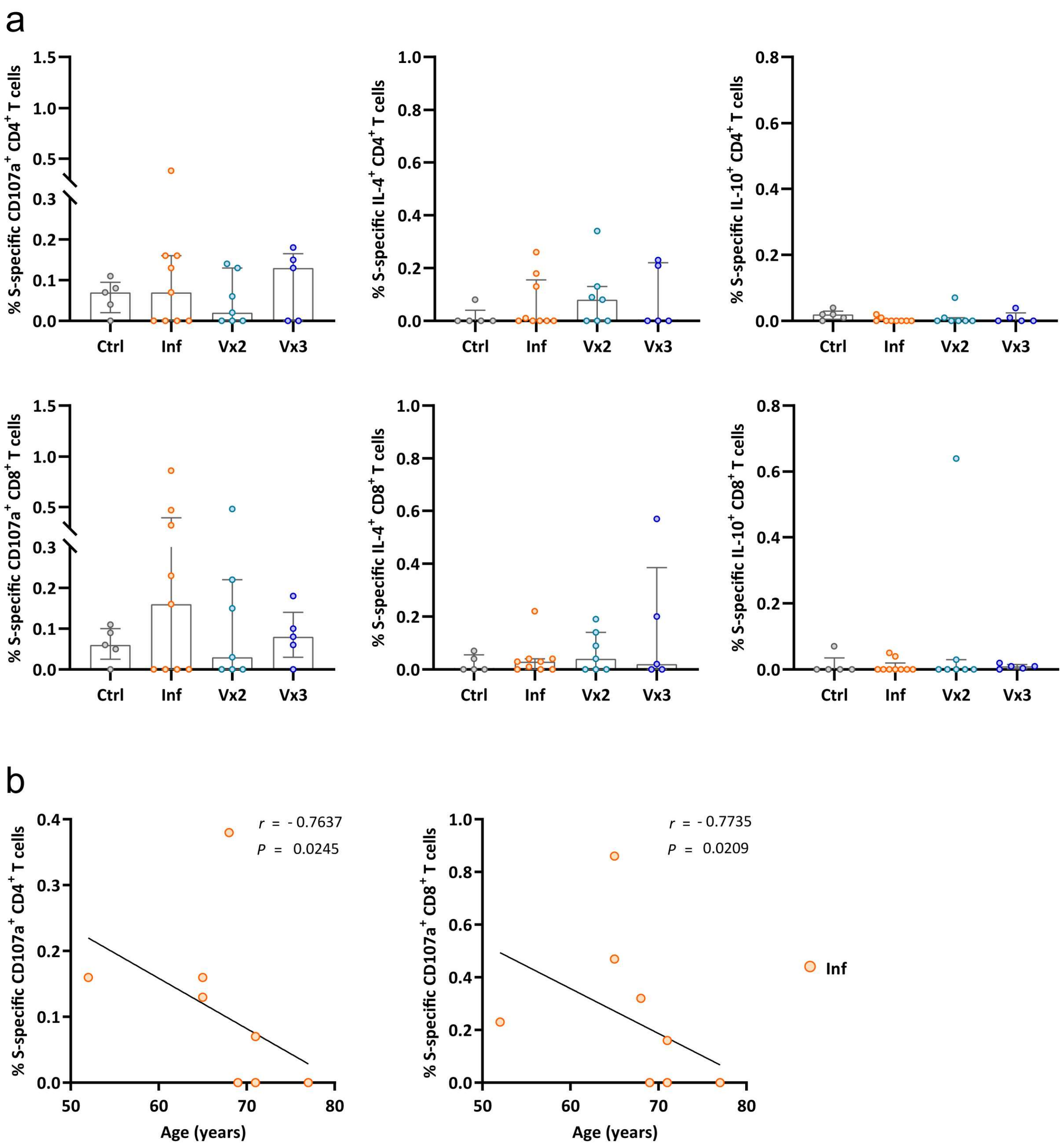

**Supplemental Figure 4. SARS-CoV-2-specific T-cell responses (CD107a, IL-4, IL-10) in the lung from convalescent and vaccinated patients.** (**a**) Comparison of the net frequency of CD107a$^+$ (left), IL-4$^+$ (middle), and IL-10$^+$ (right) cells within CD4$^+$ (upper) and CD8$^+$ (lower) T-cell subsets for each of the four groups after exposure of single-cell suspensions of lung tissue to S-peptide pools. Data in bar graphs are shown as median ± IQR, where each dot represents an individual patient for each group (Ctrl, control, n=5; Inf, convalescent infected, n=9; Vx2, vaccine 2 doses, n=7 and Vx3, vaccine 3 doses, n=5). Statistical significance was determined by Kruskal-Wallis test (with Dunn´s post-test). (**b**) Correlation between the net frequency of S-specific CD107a$^+$ cells of CD4$^+$ (left) and CD8$^+$ (right) T cells in the lung and age (Inf group). Correlations (*r* and *P* values) were assessed by Spearman test.

**Supplemental Figure 5**

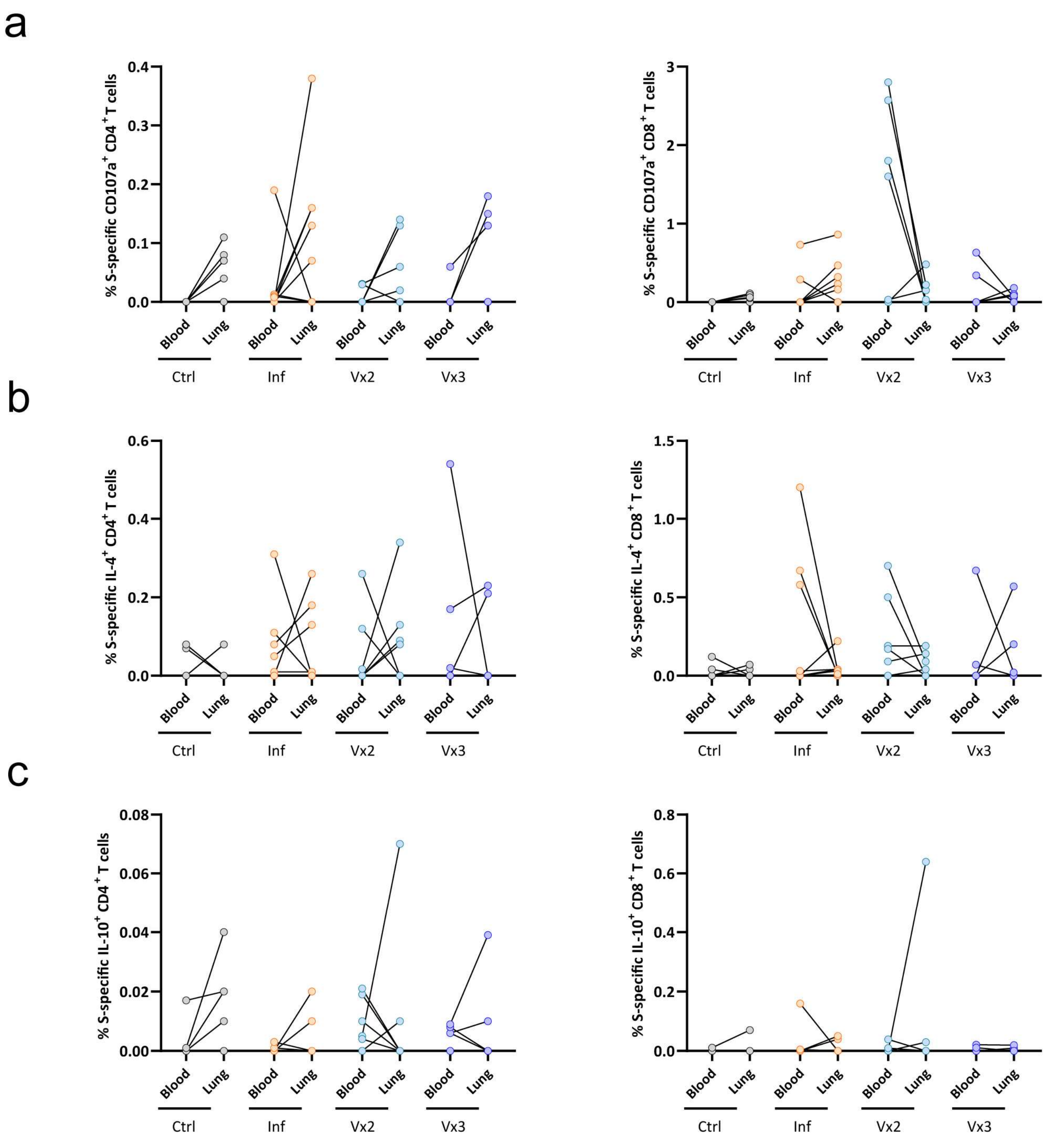

**Supplemental Figure 5. Comparison of the frequency of S-peptide specific $CD4^+$ and $CD8^+$ T cells between lung and blood.** (**a-c**) Graphs show the individual patient net frequency of $CD107a^+$ (**a**), $IL\text{-}4^+$ (**b**), and $IL\text{-}10^+$ (**c**) cells within $CD4^+$ (left) and $CD8^+$ (right) T-cell subsets of paired blood and lung samples that were exposed to S-peptide pools. Statistical significance was determined using Friedmann test (with Dunn´s post-test) for the difference between blood and lung samples within each patient group.

### Supplemental Figure 6

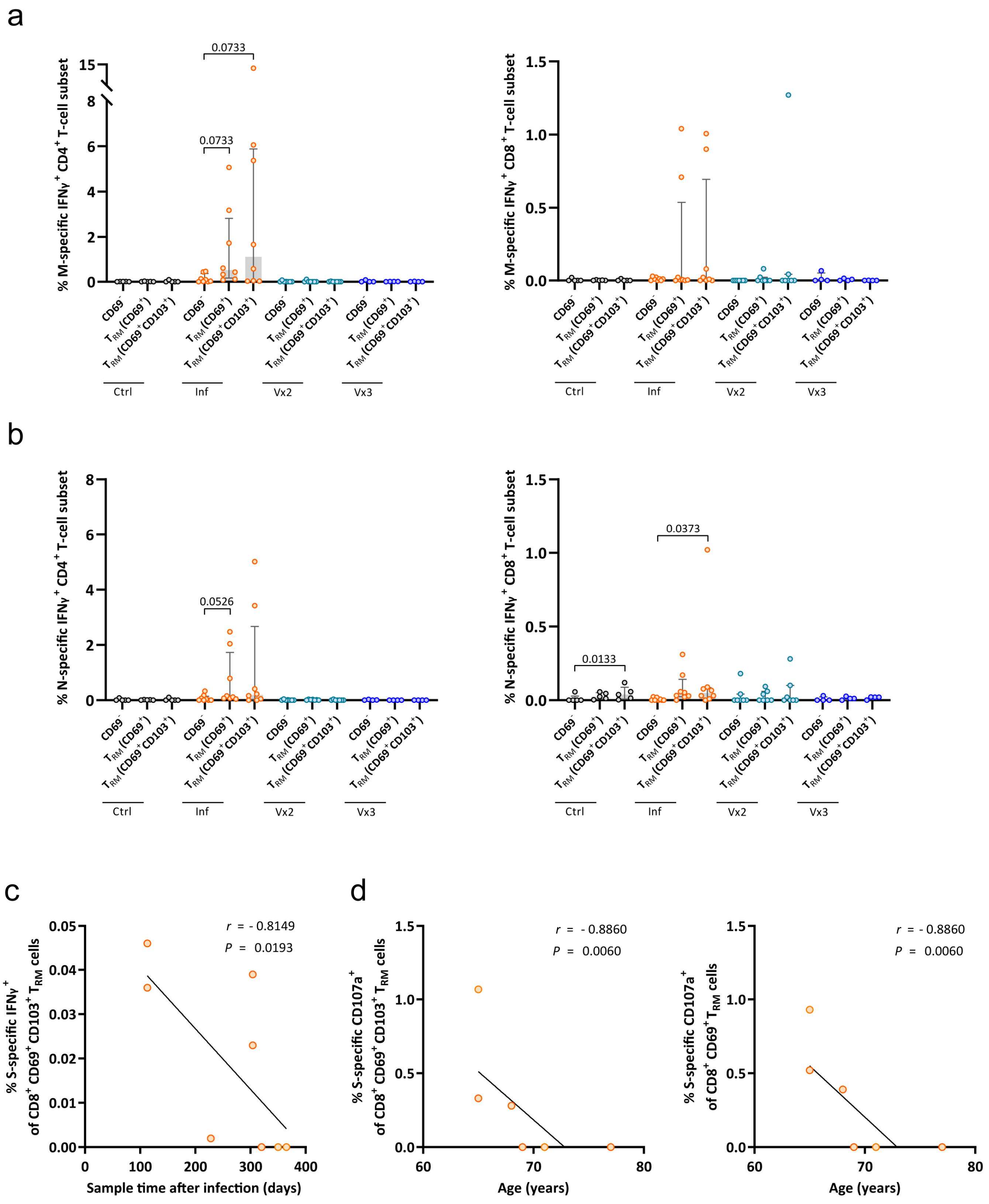

**Supplemental Figure 6. The response of $CD4^+$ and $CD8^+$ (non-) $T_{RM}$ cells against M and N peptide pools.** (**a, b**) Comparison of the net frequency of $IFN\gamma^+$ cells within three $T_{RM}$-cell subsets of $CD4^+$ (left) and $CD8^+$ (right) T cells present in the lung: $CD69^-$ non-$T_{RM}$, $CD69^+$ $T_{RM}$, and $CD69^+CD103^+$ $T_{RM}$ cells for each group after exposure to (**a**) M- and (**b**) N-peptide pools. Data in bar graphs are shown as median ± IQR, where each dot represents an individual patient for each group (Ctrl, control, n=5; Inf, convalescent infected, n=8; Vx2, vaccine 2 doses, n=7 and Vx3, vaccine 3 doses, n=4). Statistical significance was determined using Friedmann test (with Dunn´s post-test) for the difference between the cellular subsets within each patient group. (**c**) Correlation between the net frequency of lung S-specific $IFN\gamma^+$ cells within the $CD8^+$ $CD69^+$ $CD103^+$ $T_{RM}$ subset and days since confirmed infection and sampling (Inf group). (**d**) Correlation between the net frequency of S-specific $CD107a^+$ $CD8^+$ $CD69^+$ $CD103^+$ $T_{RM}$ cells or $CD8^+$ $CD69^+$ $T_{RM}$ cells and age (Inf group). Correlations (*r* and *P* values) were assessed by Spearman test.

### Supplemental Figure 7

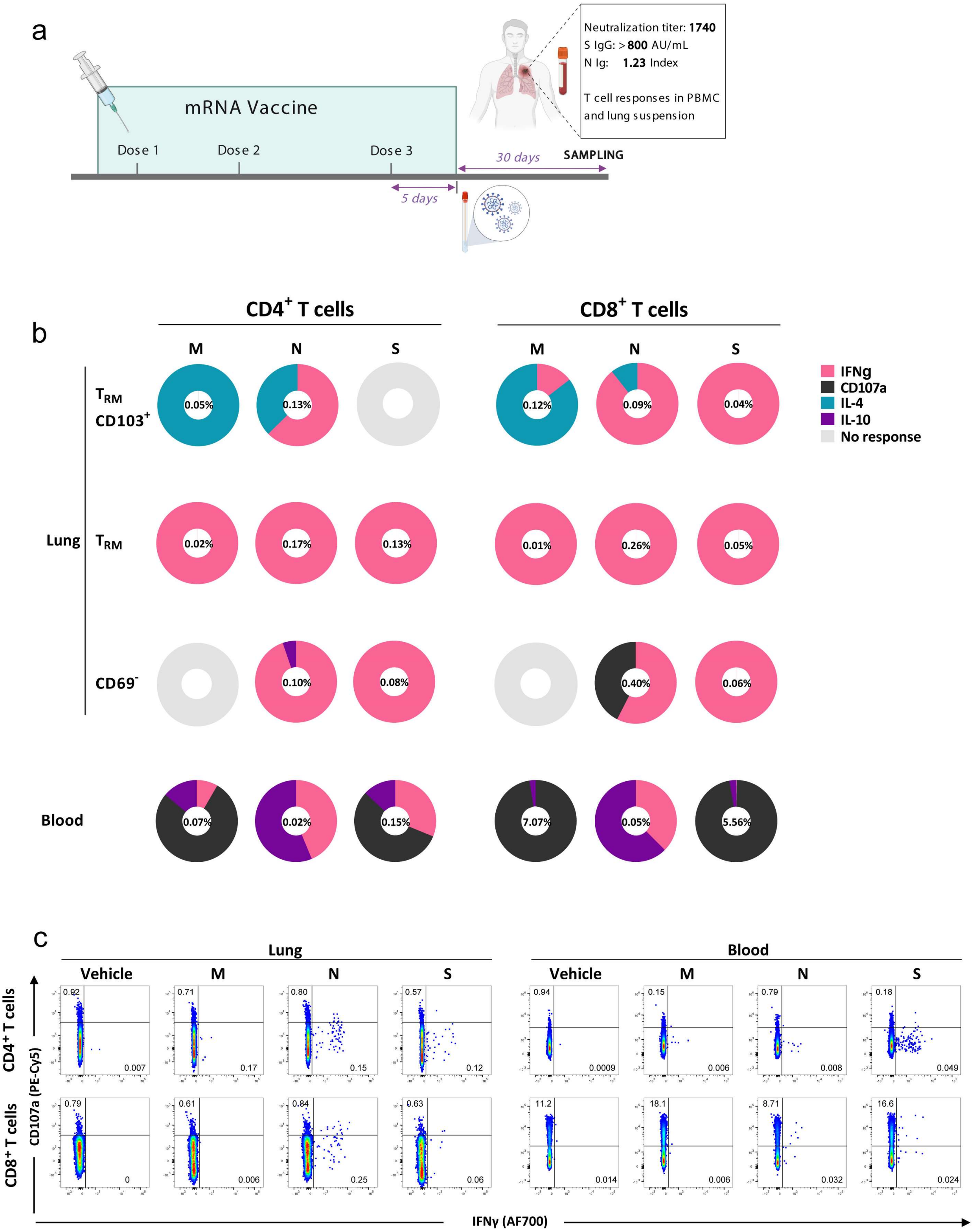

**Supplemental Figure 7. T-cell responses for patient #174.** (**a**) Timeline for patient 174, indicating vaccination against SARS-CoV-2, infection with SARS-CoV-2, and acquisition of blood and lung samples. (**b**) Donut charts displaying the net contribution of each functional marker (IFNγ, CD107a, IL-4, IL-10, or no response) to the M-, N-, and S-specific $CD4^+$ and $CD8^+$ T-cell response within the lung resident and non-resident T-cell subsets and in peripheral blood for patient 174. The frequency shown inside each donut chart represents the accumulated mean response of all functions. (**c**) Flow-cytometry plots of patient 174 showing $CD4^+$ (upper) and $CD8^+$ (lower) T cells expressing CD107a and IFNγ after exposure of lung single-cell suspensions (left) and PBMCs (right) to M-, N- and S-peptide pools or left unstimulated.

### Supplemental Figure 8

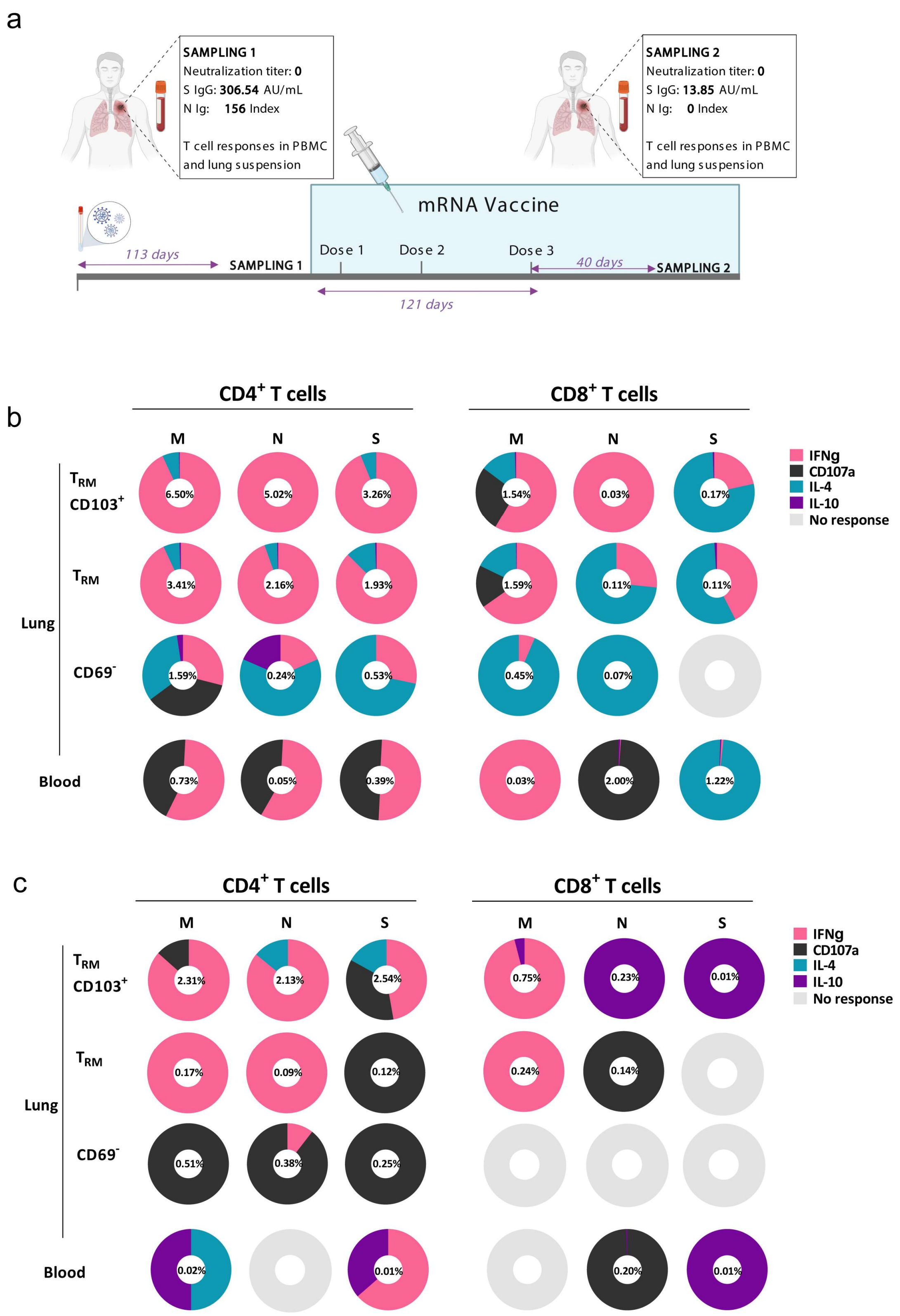

**Supplemental Figure 8. T-cell responses for patient #162, longitudinal samples.** (**a**) Timeline for patient 162 who was longitudinally sampled, first ~4 months after SARS-CoV-2 infection and then ~1 month after third dose mRNA-vaccination. (**b, c**) Donut charts displaying the net contribution of each functional marker (IFN$\gamma$, CD107a, IL-4, IL-10, or no response) to the M-, N-, and S-specific $CD4^+$ and $CD8^+$ T-cell response within the lung resident and non-resident T-cell subsets and in peripheral blood for patient 162 in the first sample (**b**) and second sample (**c**). The frequency shown inside the donut chart represents the accumulated mean response of all functions.

### Supplemental Figure 9

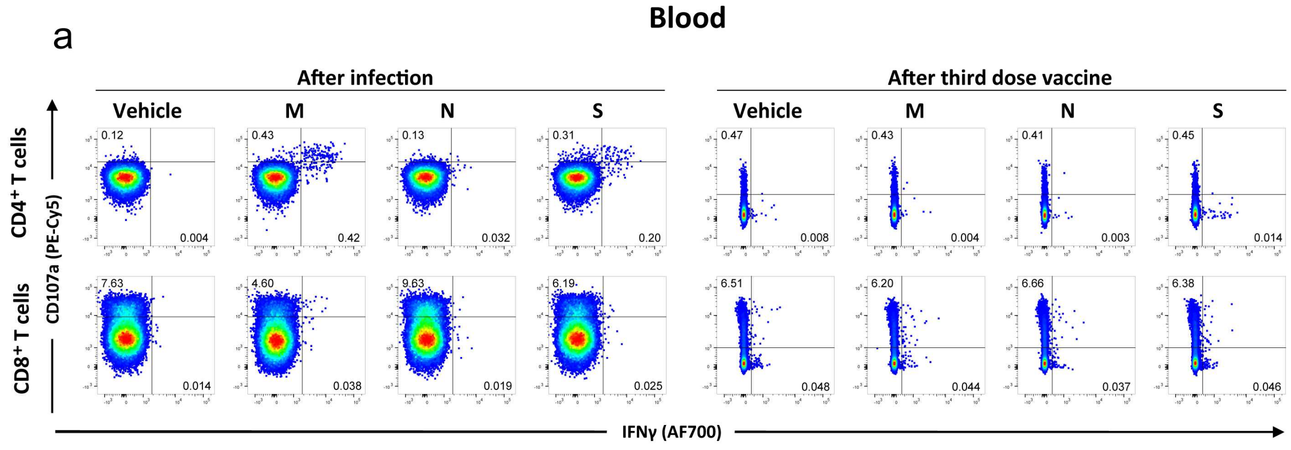


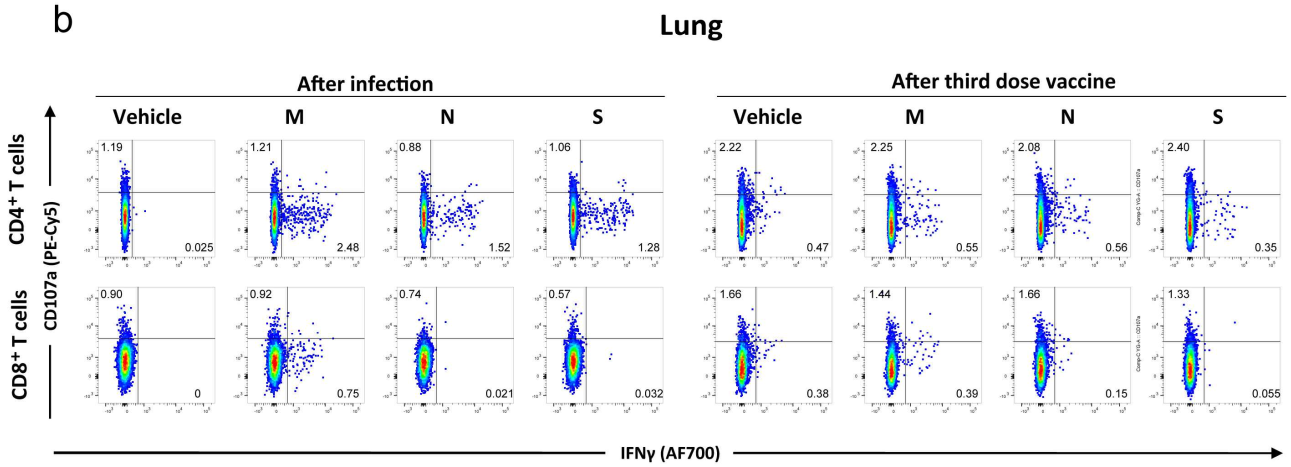


**Supplemental Figure 9. Longitudinal patterns of SARS-CoV-2-specific T-cell responses in lung and blood for patient #162.** (**a, b**) Flow-cytometry plots showing longitudinal data (left, after infection, sample 1; right, after third-dose vaccination, sample 2) of patient 162 showing $CD4^+$ (top) and $CD8^+$ (bottom) T cells expressing CD107a and IFNγ after exposure of PBMCs (**a**) and lung cell suspension (**b**) to M-, N-, and S-peptide pools or left unstimulated (vehicle).

**Supplemental Table 1. Patient characteristics**

| | **Control** n=5 | **Infected** n=9‡ | **Vx2** n=7‡ | **Vx3** n=5‡ | ***P* value** between groups |
|---|---|---|---|---|---|
| Age (Years), median [IQR] | **67** [66-74] | **69** [65-71] | **61** [58-70] | **72** [69-73] | 0.6555[a] |
| Female, n (%) | **1/5** (20%) | **1/9** (11%) | **4/7** (57%) | **3/5** (60%) | 0.0875[b] |
| Days after infection* or vaccination, median [IQR] | N/A | **304** [183-320] | **206** [184-234] | **52** [42-54] | **0.0006**[a] |
| Spike-specific IgG** (AU/mL), median [IQR] (AU/mL) | **<1.85** [1.85-1.85] | **133.1** [89.04-228.46] | **225** [118.85-248.46] | **800** [716,35-800] | **0.0122**[a] |
| Total nucleocapsid-specific Ig** (Index), median [IQR] | **0.07** [0.07-0.08] | **135** [62.1-157.5] | **0.1** [0.07-0.11] | **0.1** [0.06-0.09] | **0.0001**[a] |
| Virus neutralization titer***, median [IQR] | **0** [0-0] | **40** [19-56] | **50** [46.25-471.5] | **138.5** [67.75-665.75] | 0.0974[a] |
| Patients with SARS-CoV-2 neutralization capacity, n (%) | **0/5** (0%) | **5/7** (71%) | **5/6** (83%) | **4/4** (100%) | 0.4877[b] |
| New infection after sampling, n (%) | **0/5** (0%) | **0/9** (0%) | **1/7** (14%) | **0/5** (0%) | 0.3763[b] |

* Confirmed by RT-PCR for SARS-CoV-2

** Measured by anti-SARS-CoV-2 S and N immunoassay

*** Measured by SARS-CoV-2 neutralization assay

‡ Plasma samples were not available for every patient

N/A not available

[a] Kruskal-Wallis test with Dunn's post-test

[b] Chi-square test
